# Faster, higher, stronger – together? A bibliometric analysis of author distribution in top medical education journals

**DOI:** 10.1101/2022.03.29.22273128

**Authors:** Dawit Wondimagegn, Cynthia Whitehead, Carrie Cartmill, Eloy Rodrigues, Antónia Correia, Tiago Salessi Lins, Manuel João Costa

## Abstract

**Introduction:** Medical education and medical education research are growing industries that have become increasingly globalized. Recognition of the colonial foundations of medical education has led to a growing focus on issues of equity, absence, and marginalization. One area of absence that has been under-explored is that of published voices from low- and middle-income countries. We undertook a bibliometric analysis of five top medical education journals to determine which countries were absent and which countries were represented in prestigious first and last authorship positions.

**Methods:** Web of Science was searched for all articles and reviews published between 2012 and 2018 within *Academic Medicine*, *Medical Education*, *Advances in Health Sciences Education*, *Medical Teacher*, and *BMC Medical Education*. Country of origin was identified for first and last author of each publication, and the number of publications originating from each country were counted.

**Results:** Our analysis revealed a dominance of first and last authors from five countries: USA, Canada, United Kingdom, Netherlands, and Australia. Authors from these five countries had first or last authored 74% of publications. Of the 195 countries in the world, 53% were not represented by a single publication. There was a slight increase in the percentage of publications from outside of these five countries from 22% in 2012 to 29% in 2018.

**Conclusion:** The dominance of wealthy nations within spaces that claim to be international is a finding that requires attention. We draw upon analogies from modern Olympic sport and our own collaborative research process to show how academic publishing continues to be a colonized space that advantages those from wealthy and English-speaking countries.

**Key messages:** *What is already known on this topic:*

-Authors from a small number of high income countries are over-represented in published journal articles on medical education.

*What this study adds:*

-This study shows that almost three-quarters of first and last authorship positions in several prominent medical education journals are held by authors from only five countries: USA, Canada, UK, Netherlands, Australia.
-Authors from low- and middle-income countries, and from countries where English is not the dominant language, are under-represented in prestigious first and last authorship positions within the medical education literature.
-As a field that claims to be international in scope, perspectives from outside of these five dominant countries are under-represented, limiting the breadth of views that make up the field of medical education.

*How this study might affect research, practice or policy:*

-This study provides support for academics, academic institutions, and academic publishers in establishing policies that prioritize the inclusion of authors from low- and middle-income countries and from countries in which English is not the dominant language.
-Explicitly including descriptions of the ways research teams address potential power imbalances in research studies that involve collaboration between HIC and LMIC authors, as well as fluent English and less-fluent English speakers in English language publications may allow further development of more inclusive models of international research collaboration.

## Introduction

The global need for well-educated doctors, and other health professionals, who are able to provide high quality health care, is undisputed given the many health challenges societies face in the twenty-first century. Scaling up of health professions education opportunities has been proposed as one means of generating an educated workforce for addressing health system needs and has been a priority of the World Health Organization since the 1950s.^1, 2^ With global recognition of significant health worker shortages,^3–6^ this focus on increasing health worker output is necessary and unsurprising, leading to the creation of many new education programs worldwide.

Rizwan et al.^5, 6^ recently mapped trends in the globalization of medical education programs: India, Pakistan, China, and Brazil currently house the largest number of medical schools. While medical education programs in LMIC and transitional countries^7^ are being established to improve health human resources within home countries, they are also drawing foreign students, both from countries lacking education opportunities and from countries where student selection is highly competitive.^5^ Countries in Eastern Europe (such as Poland, Hungary, and the Czech Republic), Russia, Ukraine, and China have increasingly established English language medical education programs to attract international students.^5, 6^ Similarly, in an attempt to attract learners from Canada and the USA, medical education programs with curricula that emulate the American model have expanded within the Caribbean.^6^ As a global phenomenon, educating doctors is a growth industry.

Medical education is generally conceptualised as an academic endeavour, best achieved through well-planned delivery of science-informed education practices, tools, structures, and processes.^8^ To support this, research in medical and health professions education has become an area of increasing scholarly attention. This has led to a proliferation of academic activity, including publications, international conference attendance, and increasing diversity of professions, perspectives, disciplines, and theoretical approaches being recognized as advancing new knowledge in the field.^9^ In examining the burgeoning literature, it is important to recognise that medical education research is, by academic measures, a relatively recent area of scholarship. It first emerged as an academic field in the 1950s in North America at a time when there was an explosion of scientific medical knowledge, an influx of financial incentives to support research, and a mandate to demonstrate greater public accountability.^10^ As an area of inquiry, its development was firmly entrenched in Euroamerican healthcare and higher education structures. The colonial underpinnings of these structures is well documented, including ways that notions of academic legitimacy are based in biomedicine (which ignore traditional and Indigenous healing practices which developed in many different global contexts over millennia^11^) and are inextricably intertwined with European colonization of parts of Africa, the Americas, and Asia.^11–16^ In North America and Europe, Abraham Flexner’s 1910 report had enormous impact, entrenching legitimacy of doctor education in high status university settings, which led to the closing of non-university-based programs including most that provided training for women and Black students.^17–20^

Recognising the colonial and Flexnerian foundations of medical education provides a helpful starting point for examining issues of equity, absence, and marginalization of diverse perspectives within current structures.^21^ In recent years, explorations of representation, discrimination, harassment, silencing, and power differentials have begun to appear in medical education journals. Many are written as commentaries and perspectives pieces, providing reflections on personal experiences and theoretical explorations of ways that dominant approaches (generally white and Euroamerican-centric) constrain and limit the field.^22–30^ There are some empirical studies examining various aspects of representation within medical education, with recent attention given to gender, sociocultural, and racial equity within academic medicine’s leadership, student body, and curricula.^31–46^ There is also growing documentation of the paucity of published voices from low- and middle-income countries and non-English speaking scholars in medical education journals that position their reach as international.^56–60^ This parallels the relative absence of authors from LMIC and non-English speaking countries in leading academic journals in many other areas of academia, including health and education.^25, 47–55^

Bibliometric analyses are one way to identify imbalances, and a growing set of papers are exploring the underrepresentation of authors from outside of North America and Europe.^50, 51, 61, 62^ Maggio et al.^62^ specifically examined authorship of knowledge syntheses by country, with authors from highly ranked North American institutions being dominant. By categorizing lead authors by UN region,^63^ Buffone et al.^50^ found that the majority of authors in the medical education literature were from North America, Northern Europe, Western Europe, or Australia.

Thomas^51^ analysed authorship by country of affiliation over a two-year period, comparing medical education journals to those in education, medicine, and biomedical sciences. Examining for all authorship positions, he found that there was greater dominance of authors from the USA, United Kingdom, Canada, and Australia in medical education than in other areas. While these studies show that there is an overall dominance of authors from high-income English-speaking countries, there has not yet been a quantification of *prestigious* authorship positions by *country*.

In medical education research, first and last author positions are often considered more prestigious and desirable. For many researchers, numbers of first and last authored publications contribute to academic recognition including promotions, tenure, awards, salary support, and access to financial support for graduate students and research projects. In addition to individual academic accomplishment, regularly publishing in highly regarded journals in one’s field allows authors to engage in academic debates and shape understandings of which topics are deemed meritorious, noteworthy, and interesting. Powerful voices in these academic journals thus help to map the academic landscape, drawing boundaries and labelling worthy areas of exploration. While it is acknowledged that first and last authorship positions denote a higher level of credit for the work, Hedt-Gauthier et al.^57^ found that health research conducted in Africa, or about Africa, was less likely to have first and last authors from low- and middle-income countries (LMICs) when the publication included collaborating authors from high-income countries (HICs).

While well-established guidelines for defining what constitutes authorship exist and are endorsed by many medical journal editors,^49, 65, 66^ guidelines for how authorship positions should be distributed across authors are underdeveloped. Thatje^66^ provided rules of thumb for determining first and last authorship positions within the natural sciences, noting that disciplinary and national culture may play a role in how decisions are made. Rees et al.^67^ recently noted that while standards of authorship exist within global health research, they do not address power imbalances that exist between authors from LMIC and HIC. A recent consensus statement by Morton et al.^68^ provided guidelines for determining author order in partnerships between LMIC and HIC scholars. However, given the recency of these guidelines, it is yet to be determined whether they will be incorporated into authorship decisions amongst partnering researchers in the field of medical education.

We undertook a bibliometric analysis of five top medical education journals to determine which countries were represented in first and last authorship positions. Our aim was to provide empirical data about which countries or regions of the world were more or less prominent in the academic spaces dedicated to medical education. While recognizing that many other journals, including predominantly clinical journals, also publish medical education research, we chose to focus on journals specifically designed to publish in this area. Thomas’s^51^ previous work was able to capture articles on the topic of medical education that were published within a broad range of clinical, specialty, and disciplinary journals with scopes not exclusive to medical education research. We aimed to build upon the work of Thomas^51^ and chose to focus on journals that primarily published within the field of medical education and health professions education, as they constitute spaces where debates and critiques are intended for audiences who tend to live and breathe within the sphere of medical education. In doing so, our aim was to capture the boundaries of a field that asserts to be international in scope.

In addition to conducting this research, we recognized that the process of doing the research was itself illustrative of issues that may affect publication trends. As a collaborative research team distributed across four continents, we realised that it is was important to explicitly discuss issues of power and privilege as part of our analysis meetings. We recognised that we were working our way through specific and concrete research processes and practices in which issues of power, voice, legitimacy, and representation were ever-present. As such, we agreed to keep an explicit focus on our research decisions, processes, and practices with a view to identifying ways privilege and power were manifest and managed in the shared work. We have included description of relevant aspects of these reflections in the manuscript, in addition to including a structured statement on reflexivity, positionality and limitations.

## Materials and methods

This study was conducted in two phases. In phase one, the methodology for this study was conceptualized and designed by the three Portuguese authors (ER, AC, MJC) on this paper. In July 2018 these authors performed a search of the Web of Science database for citable items (reviews and articles) that had been published between January 1, 2012 and December 31, 2016 in the five top ranking medical education journals at that time within the categories of Education and Educational Research; Education; Education Scientific Disciplines; and Health Care Sciences & Services: *Academic Medicine, Medical Education, Advances in Health Sciences Education, Medical Teacher*, and *BMC Medical Education*. The resulting dataset was extracted into Microsoft Excel for data analysis. While Web of Science allowed for the export of author affiliations for all authors as a single data field, it was not possible to have author affiliation for first and last authors extracted separately. Consequently, there was the need to review each item and identify the country of author affiliation for first and last authors (when articles included multiple authors). Though time consuming, this manual process of identifying country of origin for each first and last author allowed for a more accurate determination than what might be expected from assigning country of origin through the use of geocoding tools or software.^51, 69^ The number of citable items (reviews and articles) for each country with affiliated first and last authors were counted: For items in which first and last author had a single country-affiliation and were from the same country, the item was counted only once. For items in which first and last authors were from different countries, or for which a first or last author had multiple country-affiliations, the item was counted the same number of times as the number of countries that were listed. Articles were counted separately for each year and for each journal. This allowed for further evaluation of longitudinal variations over the eight years of study and between publication sources. While some bibliometric studies include only a sub-sample of publications, our inclusion of all published articles allowed for an extremely accurate count of first- and last-authored publications originating from each country.^51^

Preliminary results from phase one were presented by the Portuguese team at the AMEE 2018 conference.^70^ A few years later, this paper’s co-first authors were writing a commentary for which we wanted to cite the results presented at the conference. We learned that the Portuguese-led work had not yet been able to move forward for publication (limited local research resources and extra work required for authors from a non-English first language setting), and, with approval of the Portuguese team, found resources in Canada to update the search and move manuscript writing forward. While acknowledging that the ability to advance the work was—ironically and problematically—contingent on specific high income country (HIC) resource availability, we agreed that continuing this empirical work on publishing inequities was important. In addition of two Canadians to the Portuguese team, we also embraced the opportunity to include team members from Ethiopia and Brazil, as respected colleagues and as part of a commitment to ensure representation of low and middle income country perspectives in research being done about global research imbalances.

In phase two of this project, the reconstituted team decided to update the data collection, adding years 2017 and 2018 to the dataset, while following the same methodology as the original strategy. While we were aware that journal rankings within the field of medical education had shifted since phase one of the study, we opted to limit our analysis to the five journals originally identified to ensure consistency in methodology and in recognition of the longstanding role these journals have played in shaping the field. All aspects of data extraction, assignment of country of origin for first and last authors, and counting techniques were replicated from the earlier analysis. Final analyses treated data from both phase one (years 2012-2016) and phase two (years 2017-2018) as a single dataset. Data analyses were conducted using Microsoft Excel and differences in proportions were compared using 95% confidence intervals.^71, 72^ No institutional ethics review was sought for this work since it did not involve human subjects. Neither patients nor the public were engaged in the study design, conduct, reporting, or distribution plans of the research. The datasets used and analysed during the current study have been archived as supplementary files.

## Results

5,468 articles were extracted from Web of Science and country of origin was assigned to first and last authors based on listed institutional addresses. After articles were counted multiple times to reflect multiple geographic affiliations for first and last authors, 6,173 unique items remained for subsequent analyses.

At the time of writing there were 195 countries in the world, including 193 UN Member States, the Holy See, and the State of Palestine.^73^ For these 195 countries, across all five journals and seven years of analysis, the number of first or last authored publications originating from each country ranged from zero to 1,936. Over 53% of countries were not represented by a single first or last authored publication, approximately 10% were represented by a single first or last authored publication, and under 4% were represented by more than 100 first or last authored publications (see Table 1).

**Table 1:** Number (percent) of countries by number of publications in top 5 medical education journals, 2012-2018.

| 851 | Number of first or last authored publications | Number of countries | Percent (%) |
| --- | --- | --- | --- |
|  | 0 publications | 105 | 53.85 |
|  | 1 publication | 20 | 10.26 |
|  | 2-10 publications | 35 | 17.95 |
|  | 11-50 publications | 25 | 12.82 |
|  | 51-100 publications | 8 | 4.10 |
|  | 101-493 publications | 2 | 1.03 |
|  | >493 publications | 5 | 2.56 |

The five countries with the greatest proportion of first and last authored publications included the USA with 1,936 (31.4%) publications, Canada with 878 (14.2%) publications, the UK with 721 (11.7%) publications, Netherlands with 532 (8.6%) publications, and Australia with 494 (8.0%) publications (see Table 2). Figure 1 shows the percentage of all publications with first or last authors from these five countries, and the remaining percentage (26.1%) of publications that originated from all “other” countries combined. Together, these “big five” nations of USA, Canada, UK, Netherlands, and Australia contributed 73.9% of all first and last authored publications across all journals and years of study. Figure 2 further shows the number of publications originating from each country across the globe between 2012 and 2018.

**Figure 1:**
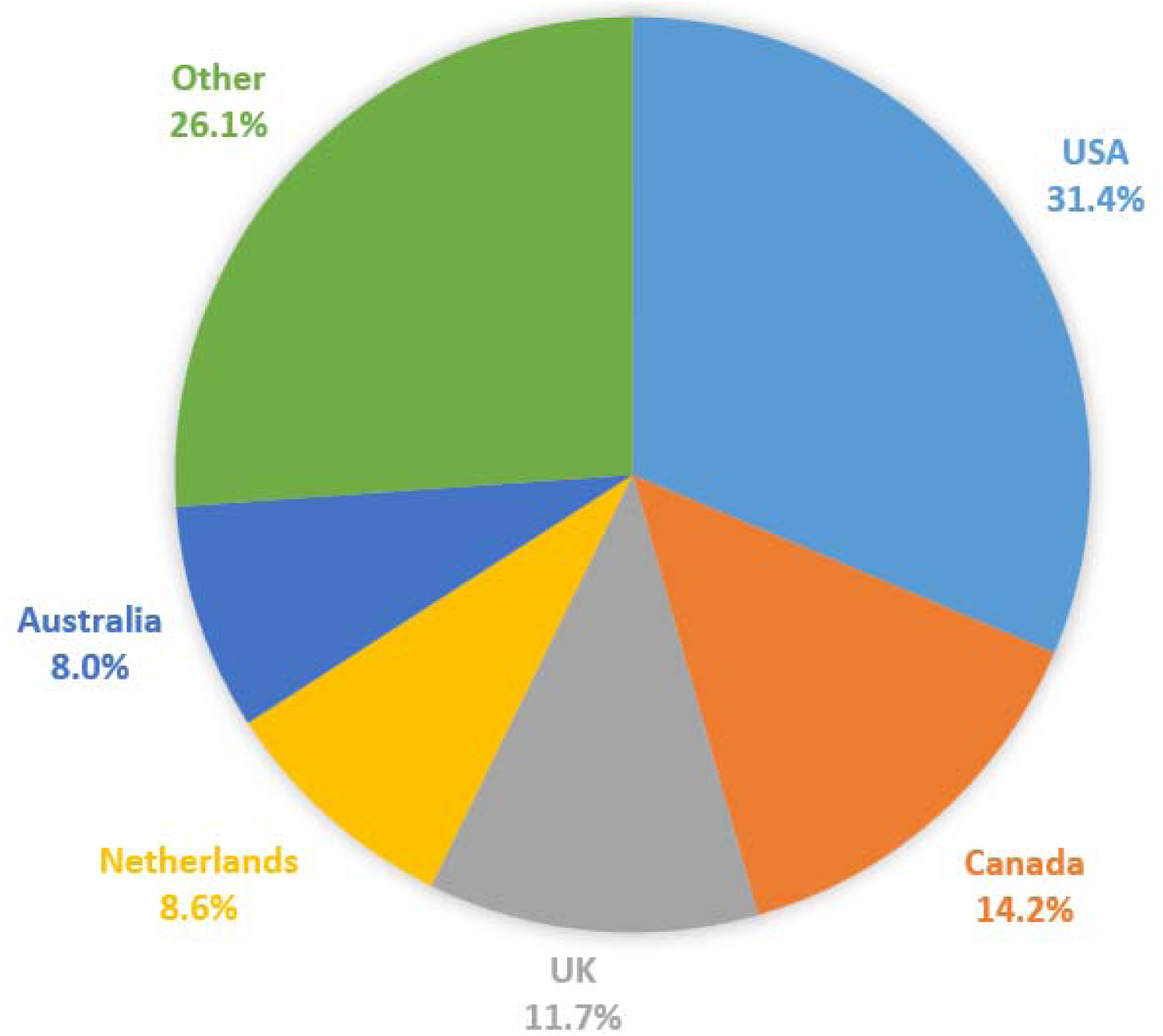
Percent first/last authored publications from USA, Canada, UK, Netherlands, Australia, all ‘other’ countries, all journals, 2012-2018.

**Figure 2:**
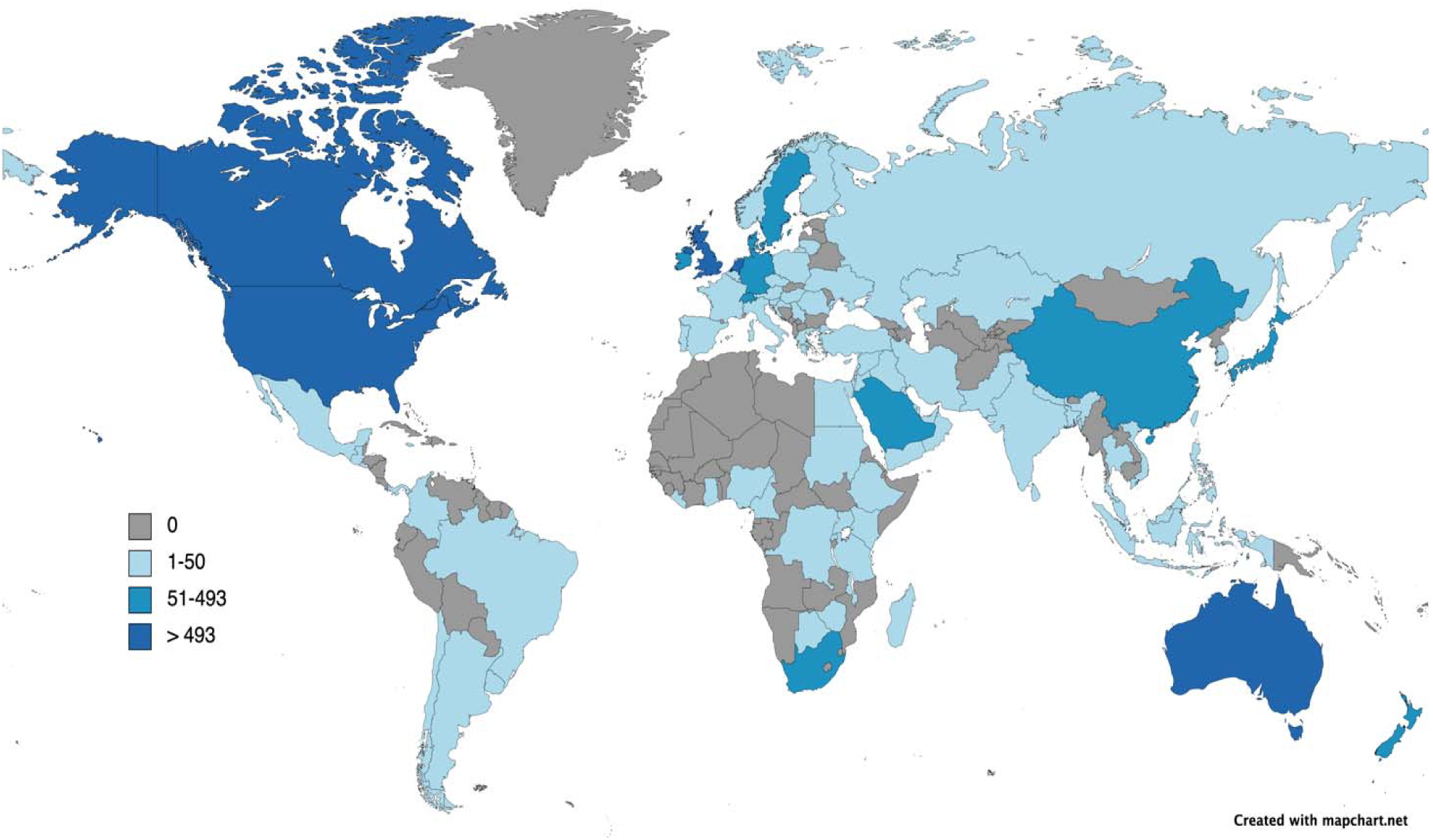
Number of first and last authored publications originating from each country, 2012-2018.

**Figure 3:**
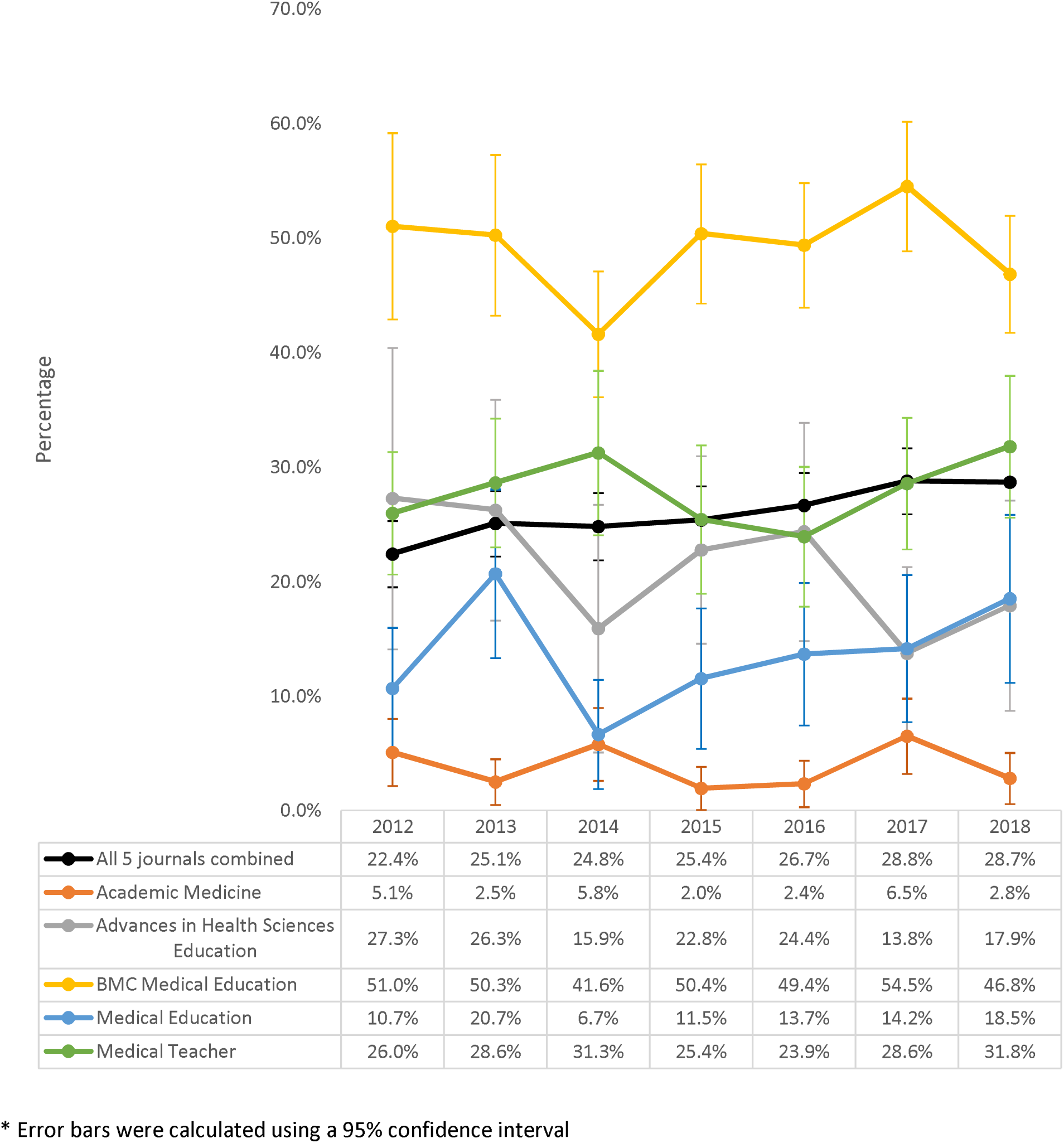
Percent* of first/last authored publications from “other” countries outside of USA, Canada, UK, Netherlands and Australia, for each journal separately and for all journals combined, from 2012 to 2018.

**Table 2:** Number and percent* of first/last authored publications from USA, Canada, UK, Netherlands, Australia, and “other” countries, by journal, 2012-2018.

|  | Acad Med | Med Educ | AHSE | Med Teach | BMC Med Educ | Total |
| --- | --- | --- | --- | --- | --- | --- |
| Impact Factor (2018) | 4.937 | 4.619 | 2.761 | 2.706 | 1.87 |  |
| USA | 1105<br>73.4%<br>(71.2-75.7%) | 164<br>20.7%<br>(17.8-23.5%) | 83<br>16.8%<br>(13.5-20.1%) | 331<br>22.3%<br>(20.2-24.4%) | 253<br>13.3%<br>(11.8-14.8%) | **1936**<br>**31.4%**<br>**(30.2-32.6%)** |
| Canada | 226<br>15.0%<br>(13.2-16.8%) | 210<br>26.5%<br>(23.4-29.5%) | 114<br>23.1%<br>(19.4-26.8%) | 188<br>12.7%<br>(11.0-14.4%) | 140<br>7.4%<br>(6.2-8.6%) | **878**<br>**14.2%**<br>**(13.3-15.1%)** |
| UK | 35<br>2.3%<br>(1.6-3.1%) | 143<br>18.0%<br>(15.3-20.7%) | 53<br>10.7%<br>(8.0-13.5%) | 275<br>18.6%<br>(16.6-20.6%) | 215<br>11.3%<br>(9.9-12.7%) | **721**<br>**11.7%**<br>**(10.9-12.5%)** |
| Netherlands | 58<br>3.9%<br>(2.9-4.8%) | 88<br>11.1%<br>(8.9-13.3%) | 98<br>19.8%<br>(16.3-23.4%) | 153<br>10.3%<br>(8.8-11.8%) | 135<br>7.1%<br>(5.9-8.3%) | **532**<br>**8.6%**<br>**(7.9-9.3%)** |
| Australia | 23<br>1.5%<br>(0.9-2.1%) | 80<br>10.1%<br>(8.0-12.2%) | 41<br>8.3%<br>(5.9-10.7%) | 121<br>8.2%<br>(6.8-9.6%) | 229<br>12.1%<br>(10.6-13.6%) | **494**<br>**8.0%**<br>**(7.3-8.7%)** |
| Other | 58<br>3.9%<br>(2.9-5.0%) | 109<br>13.7%<br>(11.3-16.1%) | 105<br>21.3%<br>(17.6-24.9%) | 414<br>27.9%<br>(25.6-30.0%) | 926<br>48.8%<br>(46.6-51.0%) | **1612**<br>**26.1%**<br>**(25.0-27.2%)** |
| **Total** | **1505** | **794** | **494** | **1482** | **1898** | **6173** |

\* Percentages were calculated with a 95% confidence interval
Acad Med: Academic Medicine
Med Educ: Medical Education
AHSE: Advances in Health Sciences Education
Med Teach: Medical Teacher
BMC Med Educ: BMC Medical Education

The inequitable presence of first and last authored publications from these “big five” Global North countries was not evenly distributed across the five medical education journals. Furthermore, there was a strong positive relationship between the impact factor (2018) of each journal and the percentage of articles that were published by first and last authors from these five dominant countries. For example, in 2018 (when our data collection ended), of the five journals studied, *Academic Medicine* had the highest impact factor at 4.937 and the highest percentage (96.1%) of articles with first and last authors from these five dominant countries (Table 2). *BMC Medical Education*, on the other hand, had the lowest impact factor at 1.87 and the lowest percentage (51.2%) of articles with first and last authors from these same countries (Table 2). Impact factor is considered one measure of prestige within academic publishing,^74^ and these results suggest that only a small number of countries are represented in first and last authorship positions within prestigious journals.

While most of our analyses combined years 2012 to 2018 together, we also looked at how the percentage of first and last authored publications from “other” countries outside of the USA, Canada, the UK, Netherlands, and Australia had changed over time. Across all journals, there was a slight increase in the percentage of first and last authored publications from “other” countries from 22.4% (95% CI 19.5-25.4%) in 2012 to 28.7% (95% CI 25.8-31.5%) in 2018. While this significant increase in publications from countries outside of the dominant five is encouraging, the overall proportion of publications from other countries remains low.

## Discussion

Our data revealed a dominance of first and last authors from five countries in five top ranked journals within the international field of medical education. Being a bibliometric analysis, the results do not provide an explanation for why there is such significant dominance by these five countries. Also, as we did not include middle authors in our research design, there may be greater presence of authors from the other 190 countries of the world than we were able to capture by looking only at first and last authorship positions. Nevertheless, the prominence of five countries in prestigious authorship positions is a finding that requires attention. While our research was not designed to identify how this dominance compared to other fields, Thomas^51^ previously found that there were greater geographic disparities within medical education research than within education, medicine, and the biological sciences. Our current research reiterated this pattern when analyzing prestigious authorship positions. Our study only included articles that had been published in the five medical education journals which had the highest impact factor at a single point in time in 2018. As a measure of journal importance, impact factor is only one metric, and one that is in constant flux. Future research might expand upon our strategy and include additional journals that publish on the topic of medical education, including journals which have experienced a significant increase in impact factor since we first devised our research strategy in 2018.

Tracking and decision-making about country assignation in bibliometric research is not always straightforward. As evidenced in previous bibliometric studies, there are many ways in which geographic origin might be determined. For the current study, we chose to double, or even triple-count publications as unique items when first or last authors claimed affiliations in multiple countries. In some instances, this led to confusion in determining whether an author had more authentic cultural connections to the Global North or the Global South. We view this as a limitation of the bibliometric nature of our study: in quantifying the geographic representation of first and last authors, we were unable to more meaningfully determine the cultural perspective from which each author was writing.

As an international research team deliberately designed to include representation from HICs and LMICs across several continents, we tried to be reflexive as we collectively advanced this project. We explicitly discussed aspects of the research process in our meetings. It was readily apparent that access to academic resources (both material resources and time) significantly shaped the research process. This included the fact that one of the Canadian authors holds a research chair that was able to provide funds for the second phase of data collection. The two Canadian authors therefore had more protected time to advance this project, which in and of itself is evidence of unearned privilege and showcases research inequities. As a team, we discussed this fact, recognized that it materially influenced the research process, while aiming to watch that it did not manifest as greater voice for these authors. We also saw this privilege as requiring greater responsibility from the Canadian authors to do the “heavy lifting” in order to move the project forward. The team also discussed the fact of English language fluency (including the genres of academic writing) and the extent to which that made the writing process more efficient for the Canadian authors to lead. We recognized that no matter how careful we were, the fact of writing in English made it easier for the Canadians to shape the words, and spent time discussing details of language in our meetings. The team decided that it was acceptable for the Canadian authors to update the preliminary research of the Portuguese team. Given both time and English language facility, the Canadian authors also led the process of manuscript writing, with the other team members making substantial edits and comments. In discussions about authorship order and thinking about contributions and ICMJE guidelines,^64^ we initially thought it might be most appropriate to have a Canadian first author and Portuguese last author based on currently accepted publishing practices and given the role that MJC had played in the original design of the study. The irony of producing a manuscript in this space with a “big five” first author was not lost on us, and we sought options to publish in other spaces that would acknowledge the significant work of authors less easily represented in current criteria. We know that HIC academics’ ability to engage in critical global work is significantly enabled by their learnings from and relationships with LMIC academic colleagues. Current authorship guidelines do not fully recognize this invisible labour on the part of LMIC academics, but as a team we decided that it was critical to incorporate these contributions into authorship decisions. As a result, we determined that co-first authorship by a LIC and HIC academic on the team was the most accurate representation of authorship. The need for such conversations further highlights the structural issues that determine who can play the sport of international academic publishing. The fact that our effort to showcase structural inequities in academia had the potential to lead to the reproduction of dominant voices is telling. In thinking about ways to increase representation in medical education research, it is clear that neither recognition of inequities nor good intentions will in and of themselves lead to structural change.

Attention to privilege and power across all aspects of a research project can lead to different conversations and different choices, with potential to help trouble assumptions that maintain HIC academic dominance. Engaging in these processes is essential; it also requires an effort that in many cases will slow research output. We believe that this “extra” work is necessary to improve knowledge creation and as a way to advance appropriate models of HIC-LMIC collaborative research practice. We also realise that this form of academic activity is not currently recognised in terms of academic metrics, and know there is more to do to find ways for such work to be valued.

The dominance of wealthy nations within spaces that claim to be international, though troubling, is not unique to medical education, the health professions, or academia in general. The prominence of the “big five” countries in medical education research brings to mind the colonization of many Global South countries during the 19^th^ Century when Global North sportsmen hunted for “big five” trophy animals: elephants, lions, rhinos, leopards, and buffalo. Symbolically, the hunting of big game in Africa and India served the purpose of solidifying the triumph of colonists over those being colonized.^75^ As a more modern sporting analogy, we are reminded of the Tokyo 2020/1 Olympic and Paralympic games, an event that represents one of the world’s largest international sporting events. Within the popular media, Tokyo 2020/1 was touted as one of the most equitable games in history. However, of the 206 nations and territories competing at the games, 98 countries had less than ten athletes participating. In contrast, the games were host to 613 athletes from the USA, 552 from Japan, 478 from Australia, 425 from Germany, 406 from China, 398 from France, 376 from Great Britain, 372 from Italy, and 370 from Canada.^76^

Rather than being a recent phenomenon, modern Olympic history, especially after World War I, has been described as paradoxically espousing universal ideals and providing an opportunity for the colonized to participate and win against the colonizers, while simultaneously reinforcing exclusionary, elitist, and racist practices.^77^ Houghton^78^ traced the history of the inclusion of Indigenous Latin American athletes and other “recently conquered” Indigenous peoples in the early Twentieth Century Olympics. In addition to participating in primitive sideshows that were aimed at showcasing the “barbaric” sporting practices of Indigenous tribes, these Indigenous peoples were also then made to compete alongside developed nations in modern Olympic events that set themselves up for ridicule, infantilization, and as a way to prove the inferiority of Global South nations.^78^ Of course, decisions about “what counts” as an Olympic sport also contribute to the dominance of wealthy nations. Sports with long colonial histories, such as football and athletics^79^ continue to be included, as do sports with more recent histories that rely on extensive and expensive sporting infrastructure, such as velodrome cycling, bobsledding, and sailboat racing. However, traditional African sports such as Nguni (stick fighting), Capoeira, donkey-racing, and Dambe boxing continue to be absent, despite recognition of their value to local peoples.^80^ We draw attention to this sporting analogy to show not only that North-South disparities are omnipresent and surreptitious, but that they are also engrained in structures and inequities that are built upon historical and colonial roots that continue to be perpetuated through international spaces, even those that aim to unite humanity and have the allure of being inclusive.

In comparing medical education research to the Olympics, we hope that looking at structures elsewhere may open up new ways of seeing a space we take for granted. Bourdieu^81^ effectively examined sport as a way to highlight that in every sphere there are philosophical underpinnings that are inherently political. Bourdieu also emphasized social spheres as spaces of conflict and struggle, including the field of science.^82^ Albert and Kleinman^83^ drew upon Bourdieu’s concepts in suggesting that it was necessary to understand how interactions that may appear to be based on cooperation may more accurately reflect domination and subordination. More recently, Martiminanakis et al.^83^ drew attention to the need to consider the inevitable knowledge politics that inform discussions about research quality and rigour within medical education. We suggest that representation in the academic literature is an area which would benefit from further exploration of the ways knowledge politics shape what is considered legitimate in these spaces. Acknowledging the skewed proportion of authors from different countries does not lead directly to solutions designed to “add” voices from LMICs without attention to the historical and colonial roots from which disparities have developed.

This leads to some potentially uncomfortable questions. For HIC researchers, it is pleasing to consider academic conversations with peers from other HICs to constitute international debates. But is it possibly a conceit to think that medical education research in its current form is truly globally relevant? To what extent is the new knowledge being shared in academic publications able to be implemented and evaluated in lower resource settings? Are important knowledges excluded from currently accepted content in medical education journals? How relevant is the content of top medical education journals in diverse contexts? How willing might HIC academics be to probe the layers of privilege that serves them well in terms of impact factor, academic promotions, and claims of international recognition? Beyond individuals, are HIC academic institutions open to questioning the structures that maintain their high international rankings? LMIC academic institutions are also driven by academic rankings so individual LMIC researchers may be encouraged to preferentially aim for international journal publications. A related issue explored by others^84, 85^ is how researchers from non-English speaking countries make choices about when to publish in “international” English-language journals versus reaching audiences in their own country and language. These are not decisions that English-language country researchers need to make, adding to the burden placed on those academics. The concept of bibliodiversity (or diversity in scholarly publishing)^86, 87^ has been adopted by Global South authors as a way of supporting the decolonization of Southern knowledge,^88^ and multilingual publishing may be one way that bibliodiversity can be achieved.^89^

## Conclusions

There have been a recent series of calls to action and incisive analyses, most prominent in critical global health spaces,^29, 47, 49, 57, 59, 90, 91^ but also in medical education journals,^30, 46, 50, 51, 62, 92^ that are contributing to a body of literature that demonstrates the ways academic publishing structures continue to privilege HIC academics. Continued conceptual and empirical work in these spaces is essential. It is also important to ensure that academics from diverse contexts are included in teams undertaking this work, and that in these collaborations close attention is paid to privilege, voice, and representation.

As a research team we believe that we must start and sustain these conversations across all aspects of our scholarly work. HIC academics, academic institutions, and academic publishers must not view the opening of cracks into privileged spaces as a way of being “nice” or as proof of benevolence. There is an ethical and moral imperative to examine and disassemble colonial structures. A recent call from the United Nations Educational, Scientific, and Cultural Organization (UNESCO)^93^ and the International Science Council^89^ compels us to consider scientific advancement a global public good to which open access is required. Eight recommendations were recently endorsed by the International Science Council,^94^ including ensuring that new scientific knowledge is accessible to all without limitations based on institutional privilege, geography, an ability to pay, or language. UNESCO also calls for more collaborative and inclusive scientific practices aimed at the achievement of the Sustainable Development Goals^95^ and reducing global inequities.

HIC also have much to learn from LMIC colleagues, and must find the humility and will to listen. As HIC countries face severe health human resource shortages, there will be learnings from LMIC colleagues who have chronically grappled with these issues. Many LMIC countries spectacularly outshone HIC countries in effectively managing successive waves of the COVID-19 pandemic^96, 97^ with far fewer resources. With growing recognition of the limits of Euroamerican biomedical approaches to healthcare, making academic space for traditional knowledges from many global contexts, as well as deep examination of the effects of colonization on health, are opportunities we must embrace. For science to be universal, it must be inclusive of a wide range of global knowledges.^94^ If scholarly medical education communities are willing to re-envisage the rules of the game to focus on ways we can be faster, higher, and stronger **together**, we may better harness the transformative aspects of education to contribute to a healthier world.

## Data Availability

All data produced in the present work have been archived as supplementary files within medRxiv.

## Acknowledgements

Not applicable.

## Contributors

DW,CW, ER, AC, MJC made substantial contributions to the conceptualization of this work. ER, AC, CC made contributions to the acquisition and analysis of the study data. All authors made contributions to the interpretation of the data. DW,CW, CC, MJC drafted this manuscript, and ER, AC, TSL critically reviewed the manuscript for intellectual content. All authors approved the final version of this manuscript, agreed to being accountable for all aspects of the work, and to investigating and resolving any questions that arise in relation to the accuracy and integrity of this work. All authors consented to having this manuscript published in *BMJ Global Health*.

## Reflexivity statement

This research was conducted by an informal partnership of authors from high-income countries, low- and middle-income countries, countries where English is the dominant language, and countries where the dominant language is other than English. Throughout the study, conversations were held during team meetings to discuss inequities that existed across team members in terms of available funds, time, and personnel for conducting and writing up the findings of this research. When deciding on an appropriate authorship order, we were guided by both the ICMJE guidelines and by the Consensus statement on measures to promote equitable authorship in the publication of research from international partnerships. We chose to take advantage of *BMJ Global Health*’s option of co-first authors. We also acknowledge that authorship guidelines can at times be used to advantage authors with greater material resources to contribute to a project, and that English language fluency greatly advantages English as first language authors in making claims to authorship position. As a team, we recognised that we were able to have ongoing and sometimes challenging conversations about the ways privilege and power needed to be examined in mundane research practices because of pre-existing trusting relationships. CW and DW have worked together for many years. TSL spent a year as a visiting scholar in Canada working with CW. Both DW and TSL have worked closely with CC on research projects. MJC, ER, and AC are Portuguese colleagues at the same institution and led the initial stage of the research project. MJC and CW had built a collegial relationship through conversations at international medical education conferences.

## Patient and public involvement

Neither patients nor the public were engaged in the study design, conduct, reporting, or distribution plans of the research.

## Patient consent for publication

Not applicable.

## Ethics approval

No institutional ethics review was sought for this work since it did not involve human subjects.

## Provenance and Peer Review

Not commissioned; externally peer reviewed.

## Data Availability

The datasets generated and analysed during this study have been archived as supplementary information files with medRxiv: https://doi.org/10.1101/2022.03.29.22273128

## Open Access

This is an open access article distributed in accordance with the Creative Commons Attribution Non Commercial (CC BY-NC 4.0) license, which permits others to distribute, remix, adapt, build upon this work non-commercially, and license their derivative works on different terms, provided the original work is properly cited, appropriate credit is given, any changes made indicated, and the use is non-commercial. See: http://creativecommons.org/licenses/by-nc/4.0/.

